# Developing a needs-based workforce plan for audiology services in England

**DOI:** 10.64898/2026.08.17.26360374

**Authors:** Saima Rajasingam, Peter MacDonald, Jagjit Sethi, Amy Taylor-Gonzalez, Amanda Hall, Imko Meyenburg

**Affiliations:** Centre for Better Living, Anglia Ruskin University, Cambridge, United Kingdom; Division of Health Economics, Anglia Ruskin University, Cambridge, U.K; NHS England, London, U.K.; Department of Audiology, Aston University, Birmingham, United Kingdom

**Keywords:** healthcare workforce, needs-based planning, audiology

## Abstract

**Background:** Internationally, healthcare workforce planning models are focussed on balancing supply and demand, rarely addressing factors such as demographic shifts and evolving health needs. For audiology services, there is a clear imperative for improved workforce planning to ensure adequate staff numbers to deliver services safely and effectively. However there is still no consensus on safe minimum staffing levels or the optimal skill mix for high-quality audiology services.

**Methods:** This research aimed to estimate the audiology health service workforce requirements in England, to meet current and projected 5 and 10 year demand for services, based on population changes and anticipated changes in demand. Informed by key stakeholders, a needs-based model was developed by (1) analysing NHS England’s National Data Collection for Audiology Services dataset to determine current workforce, (2) creating an epidemiological model to predict changes in service population over next 5 and 10 yrs, and (3) using audiology professional body endorsed estimates on East of England staff grade required per activity.

This research aimed to establish markers of quality in audiology service provision and estimate the audiology workforce requirements to meet current and projected demand for services, based on population changes and anticipated changes in demand. Following stakeholder engagement, a needs-based model was developed by (1) analysing National Data Collection for Audiology Services dataset to determine current workforce, (2) creating an epidemiological model to predict changes in service population over next 5 and 10 yrs (3) use of BAA endorsed estimates delivered in East of England on staff grade required per activity.

**Results:** The estimates for 10-year adult and paediatric audiology service whole time equivalent (WTE) safe minimum staffing levels for England (bands 2-7, current waiting times maintained) based on a population change model (Model 1), and two further models for paediatrics specifically (Model 2 and Model 3) were as follows: for adult audiology Model 1 estimates a 7.40% increase by 2035 (to 1125.18 WTE). For paediatric audiology Model 1 estimates a −6.3% (to 593.47 WTE) decrease due to underlying paediatric population decline in England, whereas the case complexities considered in Model 2 (1072.33 WTE) and Model 3 estimate a 10-year increase of 71.23% (to 1072.33 WTE) and 59.17% (to 996.82 WTE) respectively.

**Conclusions:** This is the first study to conduct a needs-based assessment of workforce requirements for UK audiology services and shows a substantial need to increase the audiology workforce. Investment in audiology workforce recruitment and training is essential to ensure that future activity levels meet population needs and that quality care is delivered. Consideration of changing demographics is required for planning future workforce specialisation. Further analysis to address workforce equity, the impact of changes in skill mix and service delivery models and local area demographics/prevalence variation is required alongside potential efficiencies.

## Background

### Needs-based planning approaches

Internationally, workforce planning in health and social care lacks consistency; models vary by country and are often shaped by local priorities. Most focus on balancing supply and demand but rarely address broader issues such as demographic shifts, evolving health needs, or long-term care pressures. Increasingly, a needs-based health workforce planning method has been adopted, but a systematic review by Lee at al., (2024) established several limitations: most models use a constant rate of disease prevalence (although disease burden is likely to vary) and few consider estimated costs in the context of economic capacity. Currently, workforce planning methods do not consider either workforce equity and inclusion (e.g., unequal career progression and disparity in opportunities for staff) or the impact of changes in skill mix and service delivery models. As health conditions grow more complex and the population ages (with multimorbidity increasingly affecting older adults as mortality rates fall), traditional staffing estimates are no longer sufficient.

### NHS Healthcare Science workforce and Audiology Services

The Healthcare Science workforce provides a range of services across the NHS (National Health Service) in England, working across over 50 different specialisms and supporting over 80% of diagnoses (Hill, 2020). Of these specialisms Audiology services provide healthcare for people with hearing loss, tinnitus and balance problems. It is distinct as one of the few specialisms within the healthcare scientist professional group who manage a whole patient pathway from diagnosis to intervention.

Hearing loss or tinnitus affects approximately 1 in 3 adults in the UK (RNID, 2026), while dizziness/balance symptoms affect approximately 1 in 3 to 5 adults (Murdin and Schilder, 2015). Permanent childhood hearing loss (deafness) has a prevalence of 1-2 per 1000 births (Wilding et al., 2023) with approximately 53,000 deaf children and young people in the UK (CRIDE, 2025) receiving regular, lifetime care from Audiology services. Otitis media with effusion (OME), the most common cause of hearing loss in children, is a highly prevalent childhood condition with 80% of children experiencing at least one episode of OME by age 4 years old (Schilder et al, 2016).

Audiologists provide diagnostics and interventions (technical, rehabilitative and psychosocial) for all these patient groups, as well as work in multi-disciplinary teams. In addition to assessment (conducting, referring for and interpreting a range of diagnostic tasks), they agree and provide tailored management plans for patients with a range of hearing, tinnitus and balance problems, drawing on a range of interventions. As Healthcare Scientists, audiologists participate in research, teaching and service improvement work.

The pipeline to become an audiologist in the UK is typically a 3-year degree course, the Audiology Practitioner Training Programme (PTP). The PTP is the entry level requirement for a band 5 position in the NHS, working with adults with hearing loss. This can be followed by the 3-year Scientist Training Programme (STP) combining clinical training with part-time Masters level study. Alternative work-based career routes including apprenticeships can also be accessed subject to funding from assistant through to BSc level. There are no minimum qualifications required to work in the audiology sub-specialties of paediatrics, tinnitus or balance. The professional body has a higher training scheme, but this is not mandatory. Much training happens “on the job” and in-house (Kingdon, 2026).

Many Healthcare Science specialisms have reported experiencing workforce shortages (IPEM 2023, ACB 2023, BAA, 2022). The consequences of these shortages have, in some cases, led to extensive failures in care. Audiology has been particularly affected. A 2022 independent review of audiology services commissioned by the Scottish Government found “significant failures” in the care of 155 children—15.7% of cases audited between 2009 and 2021 (Taylor, 2023). Early identification and effective management of hearing loss are vital for children as it can have lifelong impacts, affecting language, cognitive, emotional, educational, and social development (Kingdon, 2025). This review of audiology services in Scotland (Taylor, 2023) recommended developing a robust workforce plan to ensure safe staffing and an appropriate patient-to-staff ratio to prevent the recurrence of these incidents. It also called for a review of staff roles and skills in line with best practice and in collaboration with professional bodies. A similar review was subsequently commissioned in England and concludes that there is, among other issues, an absence of ‘coherent workforce planning’ within audiology (Kingdon, 2025). Although the British Academy of Audiology (2021, 2022) has highlighted that urgent workforce improvements are needed, there is still no consensus on safe minimum staffing levels, or the optimal staff mix to deliver high-quality audiology services. Additionally, healthcare across the NHS report experiencing psychological stress and symptoms of burnout (Weyman et al., 2024) in part due to high workload, staffing issues and emotional strain.

There is a clear imperative for improved workforce planning to ensure there are (a) adequate numbers of staff to deliver audiology services based on population need and (b) they have the right specialist training to ensure services are safe and effective. This research aimed to estimate the audiology workforce required to meet current and projected demands for services, based on population changes and anticipated changes in demand due to case complexity.

Using a needs based framework, an economic analysis was conducted to estimate the optimal workforce composition, based on population needs, current NHS workforce data (including service provider-level information on patient activity, waiting lists, and existing staffing levels), and socio-economic data from the Office for National Statistics (ONS), such as ICB (Integrated Care Board) level population data by age and other demographics or deprivation indicators, cross-referenced with existing research on hearing loss prevalence (e.g. Löhler et al., 2019; Akeroyd and Munro, 2024) to develop a socio-economic model capable of estimating future service demand at the level of individual ICB and their associated audiology departments.

## Methods

### Overview of the gap analysis

This study employed a five-step process adapted from the gap analysis process undertaken by Al-Senani et al., (2019) to quantify the workforce gap.

Initially, stakeholder engagement was conducted with 5 NHS Audiology Heads of Service and Clinical Leads with responsibility for workforce planning to identify key parameters of the workforce modelling work. Second, the NHS England’s National Data Collection for Physiological Science (Audiology)was analysed to determine the current Audiology workforce in England. Third, an epidemiological model was created to predict population changes over the next decade (assuming a constant prevalence of hearing loss). Population projections by ONS (2025) for England were used to estimate average paediatric and adult ICB and sub-ICB population changes for 2030 and 2035. 2026 ICB reorganisation were not used. Further, all projections assumed that the proportion of the population requiring hearing healthcare remains constant over time, i.e. we do not assume that there will be a future change in the current percentage of population seeking hearing healthcare in NHS audiology departments (see (Tsimpida et al., 2024). Fourth, guidelines produced by East of England Services and endorsed by the professional body (British Academy of Audiology) were used to estimate staff-to-patient ratios. Fifth, these elements were combined with future expectations on demand over 10 years. Additionally staff salary costs were estimated for each model.

### Step 1: Stakeholder engagement

An online focus group with 5 NHS Audiology Heads of Service and Clinical Leads with responsibility for workforce planning were held in July 2024. Stakeholders were consulted on the workforce modelling methodology and gave their views on key considerations. This included (1) the need to base workforce models on the requirements to meet UKAS (United Kingdom Accreditation Service) IQIPs (Improving Quality in Physiological Services) accreditation; (2) for workforce models to have some redundancy (i.e., factor in staff sickness, maternity, training capacity and the reduced productivity when training new staff); (3) the need to consider the changing nature of audiology service users (i.e., the current limited reach of services to older adults with cognitive issues and increasing numbers of children referred with social and communication difficulties); (4) the need to consider the impact of alternative service provision and pathways on workforce requirements (i.e., an audiology first pathway could reduce inappropriate and unnecessary referrals and appointments to ENT but requires a workforce with a higher skill-mix).

Points 1 & 2 have been accounted for in the following workforce estimates (by basing the model on IQIPs accredited sites only, and assuming that current staffing and activity numbers reflect sickness leave and trainee allocations) but 3 & 4 will require additional work with national leads and commissioning guidance in future iterations of this model to established likely alternative pathways and increasing access to services. Additionally point 3, highlights some limitations of a needs-based approach that solely uses prevalence data. 20.3% of the global population is estimated to have hearing loss, (Haile et al., 2019) but in England, it is estimated that only 40% of those with acknowledged hearing loss disclose this to a healthcare professional (Tsimpida et al., 2024). On that basis, the planned model would use a mixture of a needs and demand-based approach to identify requirements.

### Step 2: Current workforce estimates

In 2022 and as part of the National Physiological Science Transformation programme, NHS England began collecting data from service providers on their staffing, estates, waiting lists, and activity levels for a specified month in the year, cycling across all physiological science areas. This study used the data from the audiology data collection for June 2025 (the second collection for audiology).

### Steps 3-5: Estimating current and future workforce requirements

This study employed a structured modelling approach to estimate current workforce requirements for England. UKAS IQIPs accredited sites were chosen to build the model as stakeholders identified that obtaining IQIPs accreditation was an indicator of a quality service but tended to require more staff resourcing and a different skill mix than the staff composition of non-IQIPs accredited services (appendix 5).

Clinical activity data was usedincluding completed procedures, waiting list numbers and referrals over 6 weeks, and staffing levels), reported in June 2025’s National Physiological Science Transformation Programme data collection initiative supplemented by a BAA-approved NHS East of England guidance on role allocation and time distribution (see Appendix 3 and 4):

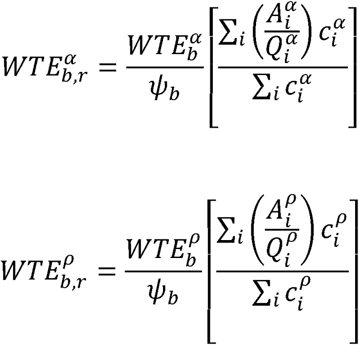

Where 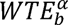 and 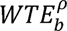 are the reported baseline *α* adult and *ρ* paediatric WTE staffing levels for AFC Band *b* respectively (here 2025), 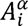 and 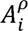 are the total adult and paediatric activity *i*, comprising 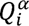 and 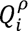 as baseline completed adult and paediatric activity *i* respectively, referrals, and waiting lists, 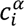 and 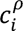 are percentage share of each adult and paediatric activity *i* for AFC Band *b* (Appendix 3 and 4), and *ψ_b_* is the contracted patient-facing time for AFC Band *b*. We finally estimated 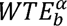 and 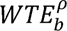 for NHS England using the ONS (2025) adult and paediatric English population numbers.

For future workforce requirements we use the above baseline model and introduce Model 1: Population Growth for adult and paediatric workforce requirements, as well as Model 2: Steady State and Model 3: Mixed Model for paediatric workforce requirements only:

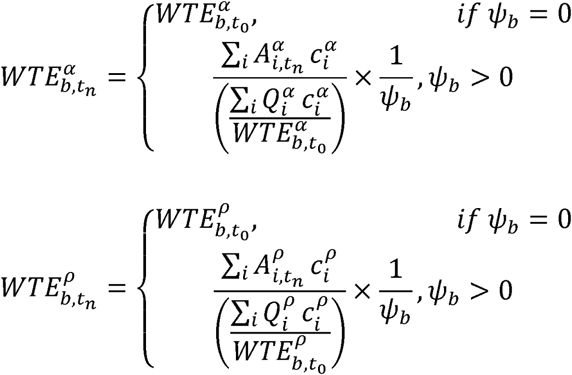

Where 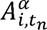 and 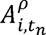 are the projected adult and paediatric activity demands at time *t*, estimated for Model 1: Population Change as:

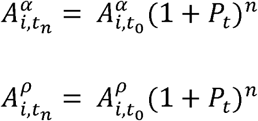

The two additional models Model 2: Steady State and Model 3: Mixed Model for paediatric activity projections only, we estimate:

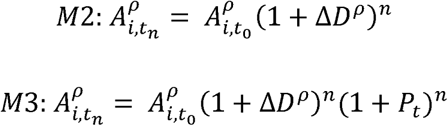

Where and *p_t_* is the population growth factor for year *t* based on ONS (2025) population estimations, and ΔD^ρ^ is a complexity adjustment for Model 2 and 3 based on the annualised increase in diagnoses and paediatric services (Bowyer et al, 2025). We use Bowyer et al.’s (2025) 6% value as a proxy for increasing service demand associated with patient complexity rather than as a direct measure of activity growth. To capture the impact of population change and case complexity adjustments, we use ΔD^ρ^ for Model 2 at 6%, while not considering growth projections with the service-specific assumptions of ΔD^ρ^ at 6%. underlying population changes, while for Model 3 we integrate ONS (2025) population growth projections with the service-specific assumptions of ΔD^ρ^ at 6%.

We also estimated 5- and 10-year total staffing costs using the current and future workforce estimations together with the mid NHS Pay Bands 2-7 annual salaries as stated from NHS Employers (2025) and adding employer national insurance (15%) and pension contributions (23.7%) based on 2026 rates. Given the absence of sufficient historical change data on those contributions to inform future projections, we assume no changes to these costs.

## Results

### Estimates of workforce requirements

Estimates for 5 and 10 year workforce requirements for adult audiology services are shown in Fig 1 for Model 1: Population Growth. Paediatric services summarised in Fig 2 include Model 2: Steady State and Model 3: Mixed Model for paediatric audiology services only to demonstrate the impact of increased demand (see appendices 6 and 7 for table format).

**Figure 1.**
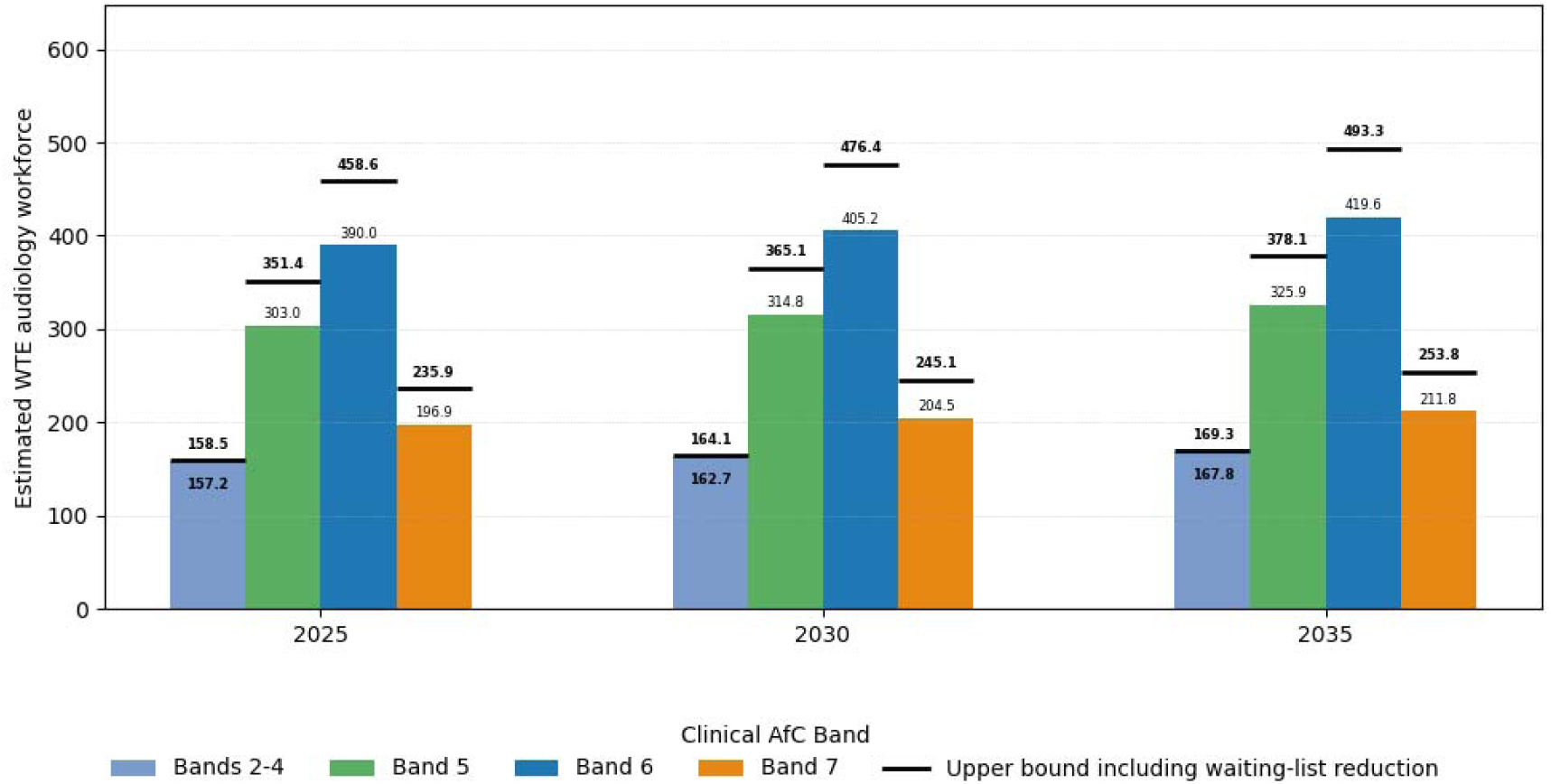
Estimated workforce requirements modelled according to population growth (Model 1) are shown as a function of Whole Time Equivalent (WTE) staff across NHS banding. Higher band numbers denote increasing in seniority and specialisation. The black lines show the upper estimates of workforce requirement, based on a reduction of the waiting times under 6 weeks (in line with NHS recommendations for waiting times for diagnostic treatment).

**Figure 2.**
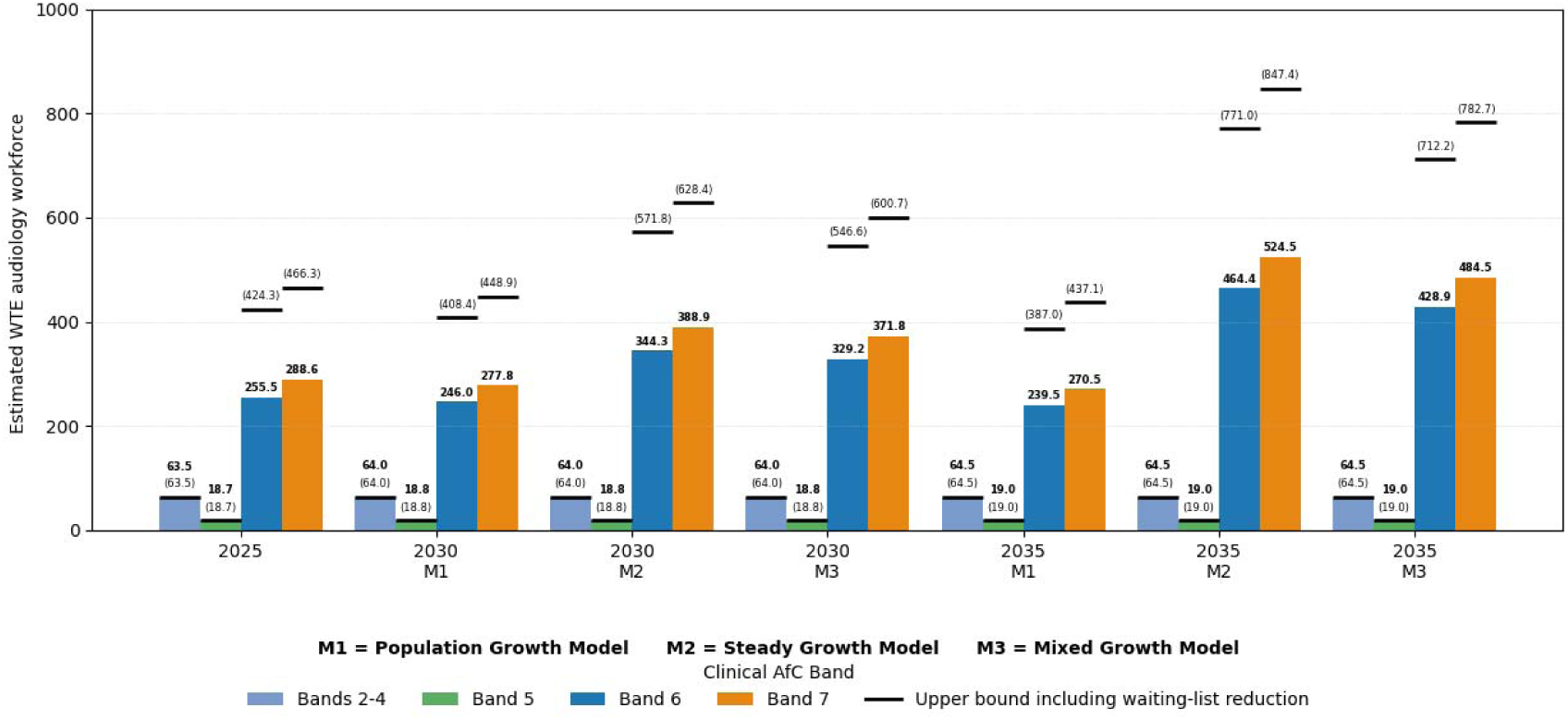
Workforce requirements shown as a function of Whole Time Equivalent (WTE) staff across NHS banding. Higher band numbers denote increasing in seniority and specialisation. The black lines show the upper estimates of workforce requirement, based on a reduction of the waiting times under 6 weeks (in line with NHS recommendations for waiting times for diagnostic treatment.

Adult audiology services are predicted to require an increase in staffing from 1047.7 to 1087.2 WTE (+3.77%) over a 5-year period and to 1125.5 (+7.40%) by 2035. As mentioned above, the ONS (2025) predicts declining population rates for under 18s across England until 2041, after which growth rates are assumed again. Consequently, our predictive Model 1 estimations for paediatric WTE staffing needs decline slightly, with minor negative growth rates of −3.15% (from 626.3 to 606.6 WTE) for 2030 and −5.24%% (from 626.3 to 593.5 WTE) in 2035 in comparison to their 2025 baseline of Z WTE.

Under Model 1, the requirement for Bands 6 and 7 are predicted to fall by 3.7% and 6.3% respectively. However, when considering the annualised increase in recorded patient complexity reported by Bowyer et al. (2025) and the likely increase in children and young people requiring audiology services has a substantial impact on the estimates with both Bands 6 and 7 increasing. Under Model 2, we see a 5-year increase to 816.0 WTE (30.29%) and a 10-year increase to 1072.4 WTE (71.23%) respectively (without waiting list reduction). For Model 3 the 5-year estimates are 783.8 WTE (25.15%) while the 10-year estimates are 996.9 WTE (59.17%) respectively. Unlike adult audiology, where the estimated increased workforce requirements are distributed equally across Bands 5-7, paediatric audiology requires a skill mix that is weighted more heavily towards Bands 6 and 7 with small 0.7% increases for both Bands 2-4 (0.5 WTE) and Band 5 (0.1 WTE) in all three models.

Title: Estimated Adult Audiology Workforce over a 5- and 10-year period.

Title: Estimated paediatric audiology workforce over a 5- and 10-year period (using population growth, steady growth and mixed growth models).

### Cost analysis

The current required workforce cost for adult audiology in IQIPS accredited hospitals is £55,873,172.70 with an 7.51% increase (£60,067,314.00) predicted by 2035.

The current required workforce costs for paediatric audiology in IQIPS accredited hospitals is £58,693,648.54. Comparing all three models, for paediatric staffing costs shows predicted changes in cost of −2.60% (£57,167,780.89) for model 1, 66.00% for model 2 (£97,433,679.86), and 60.74% for model 3 (£94,346,099.73) that the population-based estimation is the most conservative estimation of staffing costs due to the underlying predicted population decline for England until 2041 (ONS, 2025).

Total costs under Model 1: Population Growth therefore rise from £114,566,821.24 to £117,235,094.89 over a 10-year period (2.28% increase) assuming no change in waiting times. If waiting times are maintained under 6 weeks, in accordance with NHS recommendations on maximum waiting times for diagnostic services, the cost increases by 48.48% to £170,112,916.65 by 2035.

**Table 1:** Estimated annual staffing costs of the adult audiology workforce over 5- and 10-year periods under Model 1: Population Growth. Cost estimates are based on midpoint annual salaries for NHS Agenda for Change Bands 2-7 derived from the NHS Employers 2025/26 pay scales (NHS Employers, 2025), with employer National Insurance contributions (15%) and NHS pension contributions (23.7%) applied using 2026 rates. Values shown in parentheses represent the estimated costs associated with the upper workforce requirement scenario, reflecting the staffing levels required to reduce diagnostic waiting times to fewer than six weeks in line with NHS-recommended standards. Percentage differences indicate the change in estimated costs in 2030 and 2035 relative to the 2025 baseline.

| Adult WTE Costs | 2025 | 2030 | % difference | 2035 | % difference |
| --- | --- | --- | --- | --- | --- |
| Bands 2-4 | £5,990,191.12<br>(£6,042,358.39) | £6,201,834.86<br>(£6,256,034.02) | 3.53%<br>(3.54%) | £6,402,184.03<br>(£6,458,306.65) | 6.88%<br>(6.68%) |
| Band 5 | £14,071,747.63<br>(£16,321,704.36) | £14,619,835.10<br>(£16,957,426.51) | 3.89%<br>(3.89%) | £15,138,673.38<br>(£17,559,222.77) | 7.58%<br>(7.58%) |
| Band 6 | £22,083,743.95<br>(£25,964,102.20) | £22,943,894.63<br>(£26,975,390.88) | 3.89%<br>(3.89%) | £23,758,142.59<br>(£27,932,711.21) | 7.58%<br>(7.58%) |
| Band 7 | £13,727,490.00<br>(£16,447,796.79) | £14,262,168.82<br>(£17,088,430.18) | 3.89%<br>(3.89%) | £14,768,314.00<br>(£17,694,875.58) | 7.58%<br>(7.58%) |
| Total | £55,873,172.70<br>(£64,775,961.75) | £58,027,733.40<br>(£67,277,281.59) | 3.86%<br>(3.86%) | £60,067,314.00<br>(£69,645,116.21) | 7.51%<br>(7.52%) |

**Table 2:** Estimated annual staffing costs of the paediatric audiology workforce over 5- and 10-year periods (using population growth, steady growth, and mixed growth models) Estimated annual staffing costs of the paediatric audiology workforce over 5- and 10-year periods under Model 1: Population Growth, Model 2: Steady Growth, and Model 3: Mixed Growth. Cost estimates are based on midpoint annual salaries for NHS Agenda for Change Bands 2-7 derived from the NHS Employers 2025/26 pay scales (NHS Employers, 2025), with employer National Insurance contributions (15%) and NHS pension contributions (23.7%) applied using 2026 rates. Values shown in parentheses represent the estimated costs associated with the upper workforce requirement scenario, reflecting the staffing levels required to reduce diagnostic waiting times to fewer than six weeks in line with NHS-recommended standards. Percentage differences indicate the change in estimated costs in 2030 and 2035 relative to the 2025 baseline.

| Paediatric WTE Costs |  | 2025 | 2030 | % difference | 2035 | % difference |
| --- | --- | --- | --- | --- | --- | --- |
| **Model 1:**<br>**Population**<br>**Growth** | **Bands 2-4** | £3,781,529.45<br>(£3,781,529.45) | £3,798,777.75<br>(3,798,777.75) | 0.46%<br>(0.46%) | £3,817,852.34<br>(£3,817,852.34) | 0.96%<br>(0.96%) |
|  | **Band 5** | £1,776,449.02<br>(£1,776,449.02) | £1,782,551.95<br>(£1,782,551.95) | 0.34%<br>(0.34%) | £1,789,301.05<br>(£1,798,301.05) | 0.72%<br>(0.72%) |
|  | **Band 6** | £22,749,320.44<br>(£41,935,723.50) | £22,240,391.65<br>(£41,138,550.74) | -2.24%<br>(-1.90%) | £21,935,164.93<br>(£40,082,968.86) | -3.58%<br>(-4.42%) |
|  | **Band 7** | £30,386,349.63<br>(£55,632,306.34) | £29,884,050.25<br>(£55,009,434.55) | -1.65%<br>(-1.12%) | £29,625,462.56<br>(£54,777,678.18) | -2.50%<br>(-1.54%) |
|  | **Total** | £58,693,648.54<br>(£103,126,008.31) | £57,705,771.60<br>(101,729,314.99) | -1.68%<br>(-1.35%) | £57,167,780.89<br>(£100,467,800.44) | -2.60%<br>(-2.58%) |
| **Model 2:**<br>**Steady**<br>**Growth** | **Bands 2-4** | £3,781,529.45<br>(£3,781,529.45) | £3,798,777.75<br>(£3,798,777.75) | 0.46%<br>(0.46%) | £3,817,852.34<br>(“3,817,852.34) | 0.96%<br>(0.96%) |
|  | **Band 5** | £1,776,449.02<br>(£1,776,449.02) | £1,782,551.95<br>(£1,782,551.95) | 0.34%<br>(0.34) | £1,789,301.05<br>(£1,789,301.05) | 0.72%<br>(0.72%) |
|  | **Band 6** | £22,749,320.44<br>(£41,935,723.50) | £29,978,651.13<br>(£54,995,874.43) | 31.78%<br>(31.14%) | £39,590,935.75<br>(£72,295,523.83) | 74.03%<br>(72.40%) |
|  | **Band 7** | £30,386,349.63<br>(£55,632,306.34) | £39,788,434.33<br>(£72,309,800.97) | 30.94%<br>(29.98%) | £52,235,590.72<br>(£94,256,365.44) | 71.90%<br>(69.43%) |
|  | **Total** | £58,693,648.54<br>(£103,126,008.31) | £75,348,415.16<br>(£132,887,005.10) | 28.38%<br>(28.86%) | £97,433,679.86<br>(£172,159,042.66) | 66.00%<br>(66.94%) |
| **Model 3:**<br>**Mixed**<br>**Growth** | **Bands 2-4** | £3,781,529.45<br>(£3,781,529.45) | £3,798,777.75<br>(£3,798,777.75) | 0.46%<br>(0.46%) | £3,817,852.34<br>(£3,817,852.34) | 0.96%<br>(0.96%) |
|  | **Band 5** | £1,776,449.02<br>(£1,776,449.02) | £1,782,551.95<br>(£1,782,551.95) | 0.34%<br>(0.34%) | £1,789,301.05<br>(£1,789,301.05) | 0.72%<br>(0.72%) |
|  | **Band 6** | £22,749,320.44<br>(£41,935,723.50) | £29,233,591.45<br>(£53,864,926.06) | 28.50%<br>(28.45%) | £37,984,001.94<br>(£69,954,292.27) | 66.97%<br>(66.81%) |
|  | **Band 7** | £30,386,349.63<br>(£55,632,306.34) | £39,054,603.30<br>(£71,463,498.03) | 28.53%<br>(28.46%) | £50,754,944.39<br>(£92,817,795.96) | 67.03%<br>(66.84%) |
|  | **Total** | £58,693,648.54<br>(£103,126,008.31) | £73,869,524.45<br>(£130,909,753.79) | 25.86%<br>(£26.94%) | £94,346,099.73<br>(£168,379,241.62) | 60.74%<br>(63.28%) |

## Discussion

This is the first study to conduct a needs-based assessment of workforce needs for audiology services and uses the good practice reporting guidelines or health workforce projection models (Lee et al., 2024). Although estimates of audiology workforce requirements exists, these are often accounted for within the “ear and hearing professional categories” (Kamenov et al., 2021) and even when specifically noted, focus on existing workforce rather than activity levels (Pillay et al., 2020, Windmill & Freeman, 2013) which means that any shortfalls in current provision are not accounted for. In some cases, only hearing loss has been considered, although audiologists also work in diagnosis and rehabilitation of balance disorders (Garuccio J et al., 2025). Additionally, this model provides data stratified according to seniority and specialisation (banding) providing an understanding of the required skill-mix and supporting discussions on staff progression.

We have identified that the current adult audiology workforce will need to grow by approximately 7% over the next 10 years (if current waiting times over 6 weeks are maintained), primarily driven by an increase in aging of the population. The most striking finding is the projected changes required for the paediatric audiology workforce, taking account of the increase in complexity of cases, requiring a 59 to 71% increase (model dependent, current waiting lists maintained). These findings have significant implications for audiology workforce planning, specifically the paediatric audiology workforce.

The model estimates should be considered in the context of training requirements and staff development. The paediatric model in particular estimates greater needs for staff at higher band or specialisation levels, reflecting the complexity of the work and additional training requirements. However, the staff required in 2035 will require training under supervision within clinical requirements in addition to completion of MSc level training and are likely to be hired at Band 5 level and trained up over this period. This will require release of current staff at Band 6 and 7 level to provide this clinical supervision and recruitment of additional staff at Band 5 level and below that is not reflected in the estimates currently. This substantial commitment is likely also to require efficiencies in service delivery. These efficiencies should be considered together with training needs and alternative service delivery models that better meet the needs of service users.

Given the timelines required to train paediatric audiologists, action is required now to increase capacity for the next 5 to 10 years. Considering should be given in particular to delivering appropriate higher-level training and capability for experienced staff to develop within the paediatric specialty, noting the increased requirement to work with high complexity caseloads. This will require staff to have access and be supported for within-work professional development opportunities. It will also require support and retention of experienced staff and clinical educators to mentor, train and supervise, in a context where services are already experiencing staff shortages; over half of UK audiology services report being unable to fill vacancies (Audiology World News, 2023). It is also relevant to question whether this increase in paediatric workforce is feasible and whether there are other solutions that can be applied, in terms of the nature of the work itself.

The workforce model stages were broadly based on the stages outlined by Al-Senani et al., (2019), but differed in the following key ways; firstly it did not outline novel interventions or pathways to estimate staff requirements as the stakeholder group discussions identified multiple potential approaches. The next version of this model should and will consider these novel interventions or pathways in more detail. Secondly, in the use of stakeholder engagement which prioritised the need to include indicators for quality in service provision.

This additional step was critical in ensuring the model provided safe estimates rather than estimates based solely on current workforce levels across all sites. Most workforce models rarely consider quality assurance of services, but the risks with current audiology service provision identified by the British Academy of Audiology (2021, 2022) and Kingdon (2025) highlight the need for workforce planning models to include measures of safety and quality.

Although the National Data Collection for Audiology Services dataset collects data from NHS audiology services, some audiology services are provided by Any Qualified Provider in the UK, which includes private and third sector providers in addition to NHS. They are not required to report any data on staffing or activity levels to the National Data Collection for Audiology Services, so the workforce predications and cost estimates exclude these providers. Note that currently all paediatric audiology services currently are solely provided by NHS providers.

Amongst adults, regional variability in the prevalence of hearing loss has not been considered within these models and will impact on the caseload for services across different regions of the UK. Analysis of self-reported hearing loss from the English Longitudinal Study of Ageing dataset demonstrates different prevalence across UK regions (Tsimpida et al., 2020), particularly areas with higher levels of deprivation. More detailed hearing and balance prevalence data with consideration of the impact of socio-economic deprivation would also provide a better understanding of workforce need nationally.

Perhaps the biggest challenge facing audiology services is balancing the waiting times with the need to train staff. Across all Physiological Science specialisms the scientist acquires the diagnostic data, analyses and reports, differing from other areas such as imaging and pathology that have medical doctors leading, analysing and reporting their diagnostic data and scientists mostly acquiring and running the laboratory procedures. Analysis of the current workforce reveals substantial numbers of service users waiting for longer than 6-weeks for an appointment leading to large differences in the predicted workforce need based on maintaining current waiting times or reducing waits to NHS recommended levels, adding to the challenges in training or upskilling new or existing staff. It is unclear what the impact of maintaining current waiting times would be on service users or how many will experience long-term consequences of failure to treat in a timely manner. Accordingly, the models do not account for the impact of complications from untreated ear-related conditions on future caseloads or complexity of case mix.

Another limitation of this study is that it does not consider the impact of equipment or training on predicted costs. The economic costing included here is based solely on salaried time for staff undertaking clinical activities. It does not consider the cost of replacing equipment or obtaining additional clinic rooms and supplies for additional staff and clinics. Supporting staff development is likely to require a reduction in clinical time for those staff undertaking the training and for those who may be providing clinical supervision, which may either result in a drop in activity levels or involve the hiring of additional staff (permanent or temporary) to maintain activity levels, with resultant cost implications.

These models did not explore workforce equity and inclusion (particularly the underrepresentation of minority or marginalised groups at leadership levels and their experiences of working in healthcare) or the impact of changes in skill mix and service delivery models. Future models should consider inequity in workforce composition and access to training, in particular workforce equity within leadership structures (Santric Milicevic et al., 2024).

## Conclusions

Overall, the estimated 10-year audiology workforce safe staffing levels for England (total WTE numbers across banding), based on our three models are adult Model 1 = 1125.18, and for paediatric Model 1=593.47, Model 2 = 1072.33, and Model 3 = 996.82. Investment in workforce recruitment and training is essential in not only bridging the current workforce gap but also ensuring that future activity levels meet population needs. Consideration of changing demographics is required for planning future workforce specialisation. Given the increasing demand, innovative new service delivery models are required to address these workforce gaps more effectively and ensure staff can access development opportunities. Inclusion of Any Qualified Provider data in the NHS National Data Collection for Audiology Services is recommended to ensure that future model predictions are realistic.

## Data Availability

The datasets used and/or analysed during the current study are available from the National Physiological Sciences Transformation Programme on reasonable request. NHS staff can access directly here National Physiological Science Data Collection Dashboard.
The population datasets used here can be accessed from the Office of National Statistics at The datasets used and/or analysed during the current study are available from the National Physiological Sciences Transformation Programme on reasonable request. NHS staff can access directly here National Physiological Science Data Collection Dashboard.
The population datasets used here can be accessed from the Office of National Statistics at https://www.ons.gov.uk/

https://www.ons.gov.uk/

https://gbr01.safelinks.protection.outlook.com/?url=https%3A%2F%2Fapp.powerbi.com%2FRedirect%3Faction%3DOpenApp%26appId%3Dbc57ec16-c243-465a-a573-f274c27c529c%26ctid%3D37c354b2-85b0-47f5-b222-07b48d774ee3%26experience%3Dpower-bi&data=05%7C02%7Cclare.warriner%40nhs.net%7C7a2aa317ca2a4c9f360308dd5ca53f3a%7C37c354b285b047f5b22207b48d774ee3%7C0%7C0%7C638768586970961213%7CUnknown%7CTWFpbGZsb3d8eyJFbXB0eU1hcGkiOnRydWUsIlYiOiIwLjAuMDAwMCIsIlAiOiJXaW4zMiIsIkFOIjoiTWFpbCIsIldUIjoyfQ%3D%3D%7C0%7C%7C%7C&sdata=fc7MdlBVExdNwlvtD%2FTGZ5%2BLe8gldvcOomuJ0XCQZUg%3D&reserved=0

## List of abbreviations

BAA: British Academy of Audiology
ICB: Integrated Care Board
IQIPs: Improving Quality in Physiological Services
NHS: National Health Service
ONS: Office for National Statistics
PTP: Practitioner Training Programme
STP: Scientist Training Programme
UKAS: United Kingdom Accreditation Service
WTE: Whole Time Equivalent

## Ethics approval and consent to participate

Ethical approval for the stakeholder groups was obtained from and amended through the Economics, Finance and Law School Research Ethics Panel (Ethics applications ETH2425-3100, ETH2425-7040, and ETH2526-4274) in the Faculty of Business and Law at Anglia Ruskin University.

## Availability of data and materials

The datasets used and/or analysed during the current study are available from the National Physiological Services Transformation Programme on reasonable request. NHS staff can access directly here *<u>National Physiological Science Data Collection Dashboard</u>*.

The population datasets used here can be accessed from the Office of National Statistics at https://www.ons.gov.uk/

## Authors’ contributions

IM, SR and JS developed and designed the study and models. AT-G supported with data access and advised on model development. IM and PM conducted the data analysis and mathematical model development. SR undertook the stakeholder engagement and analysis. SR and IM drafted the paper. All authors contributed to the drafting and editing of the manuscript.

## Acknowledgements

The authors would like to express thanks to the National Physiological Services Transformation Programme for providing access to the National Data Collection for Audiology Services for development of this model, along with Victoria Parfect and the East of England Audiology Network and the British Academy of Audiology for development and approval of estimates of patient facing time and activities undertaken according to banding.

## Appendix 1 Good practice reporting guideline for health workforce projection models mapped against this paper*-taken from Lee et al., (2024)*

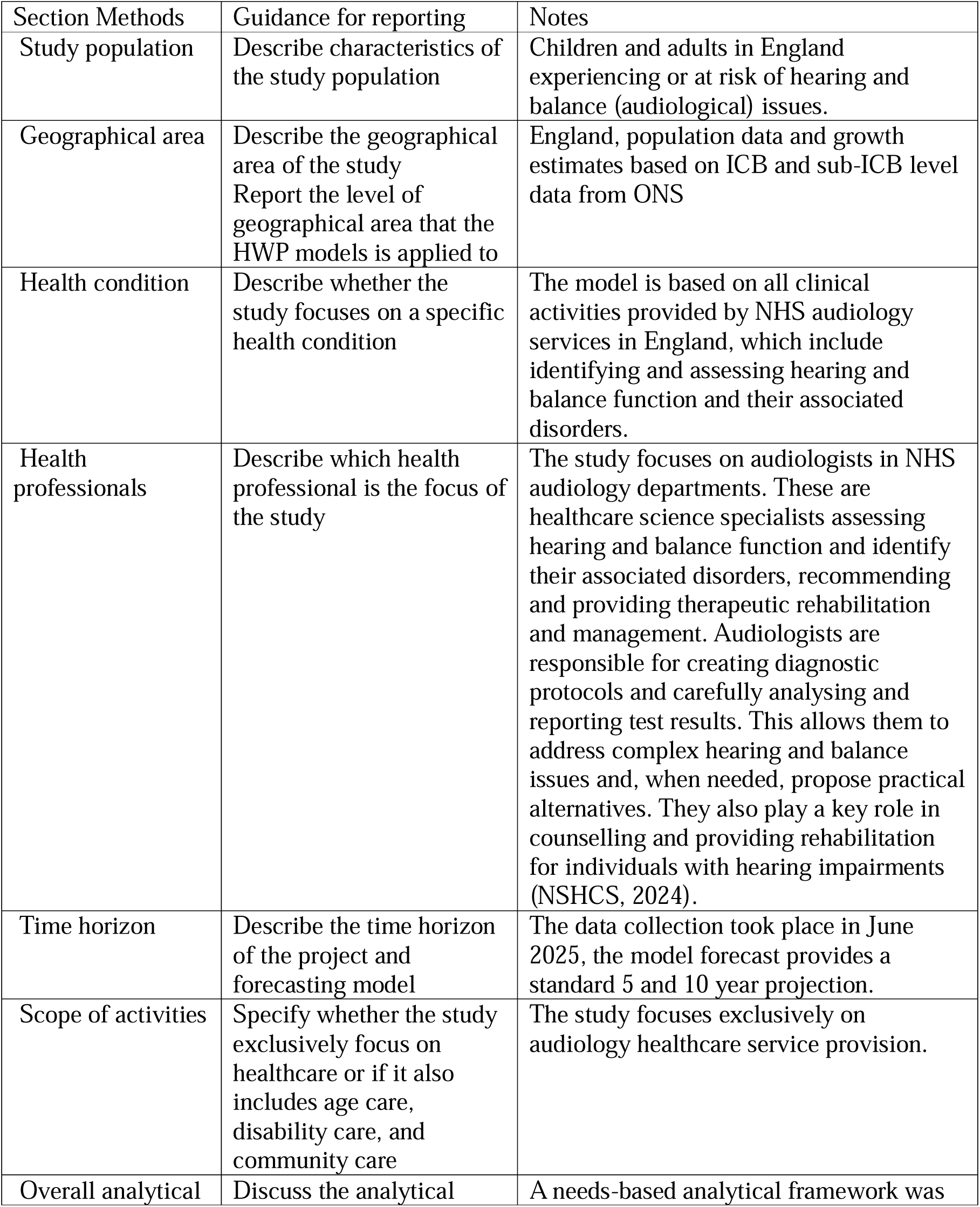
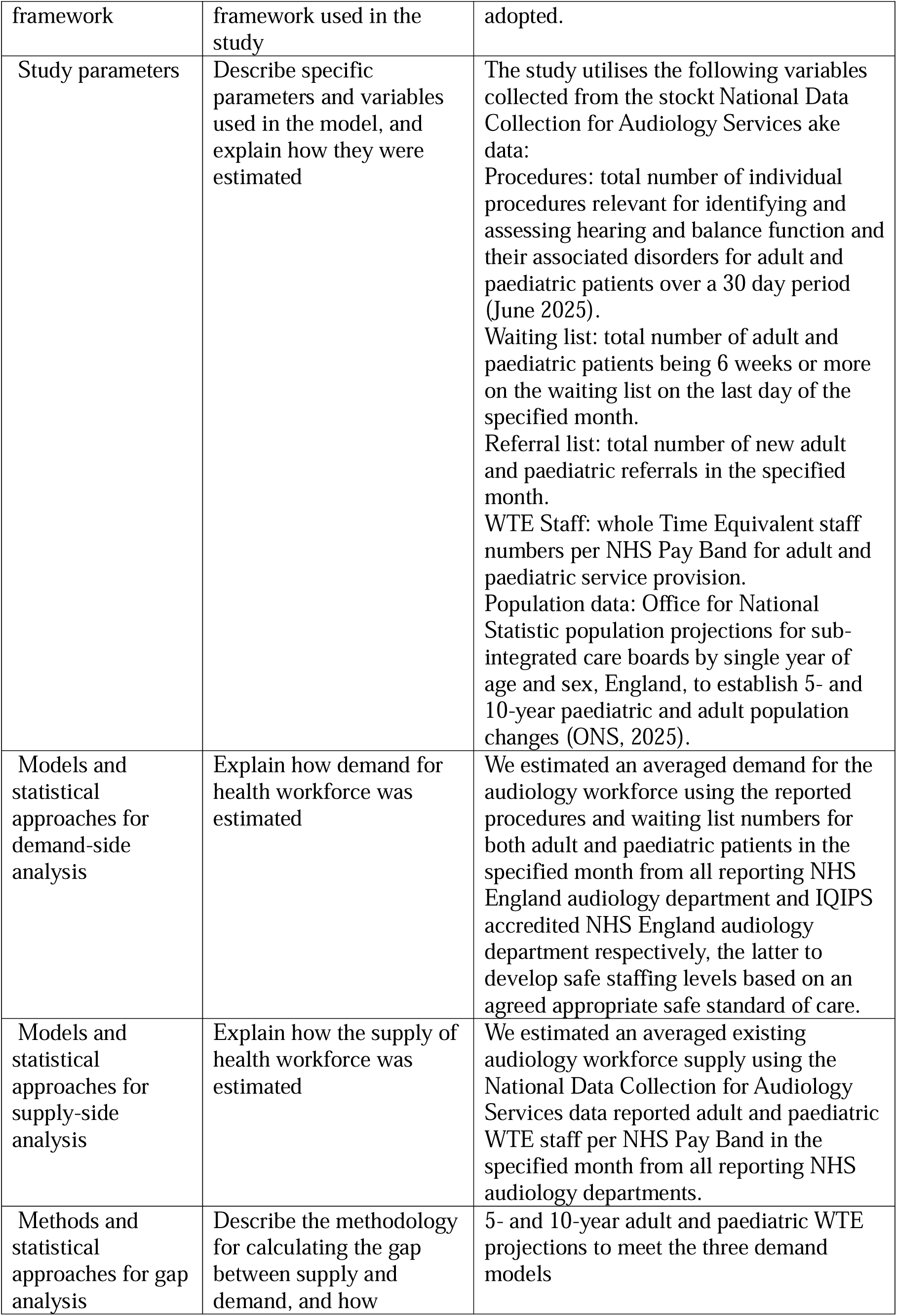
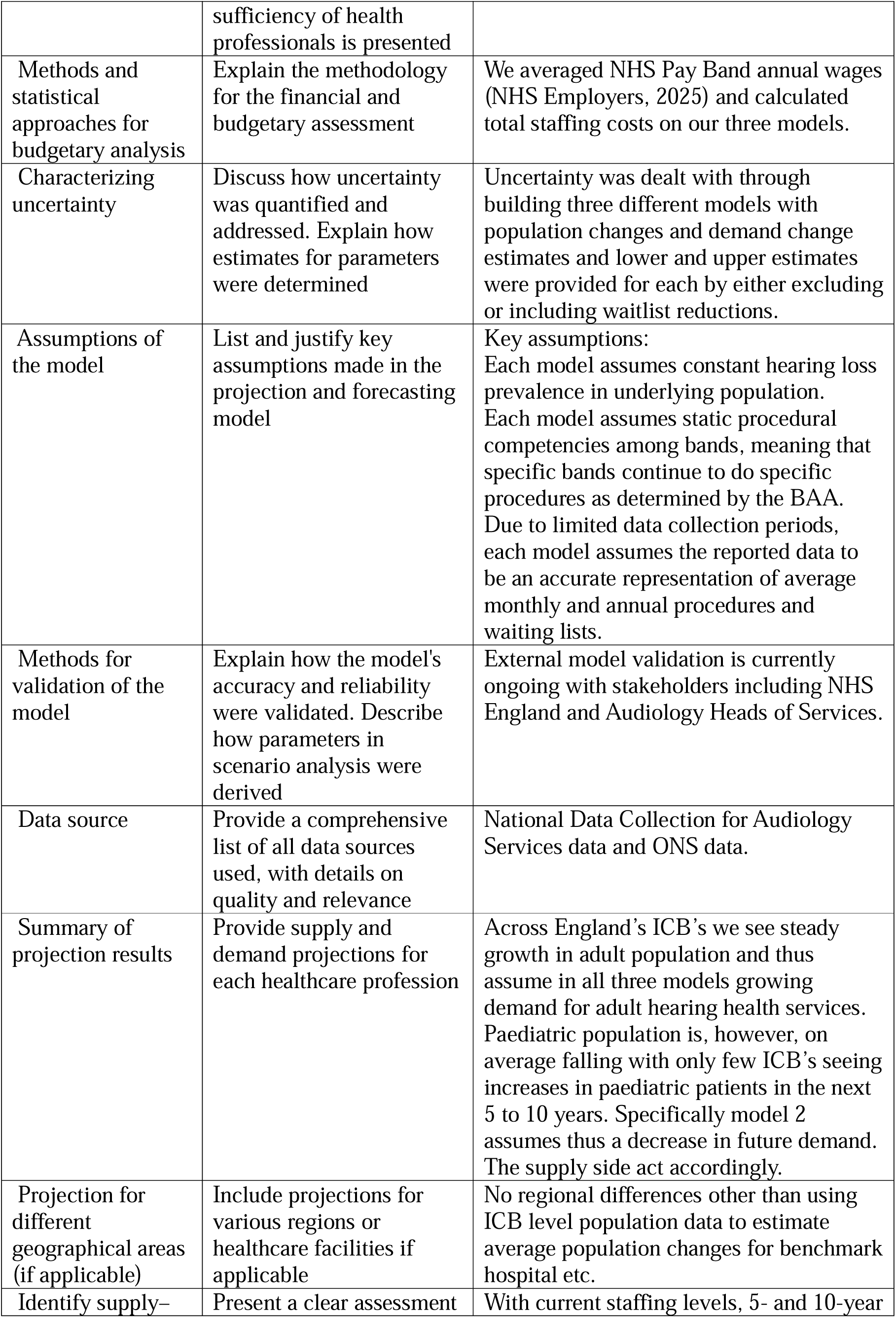
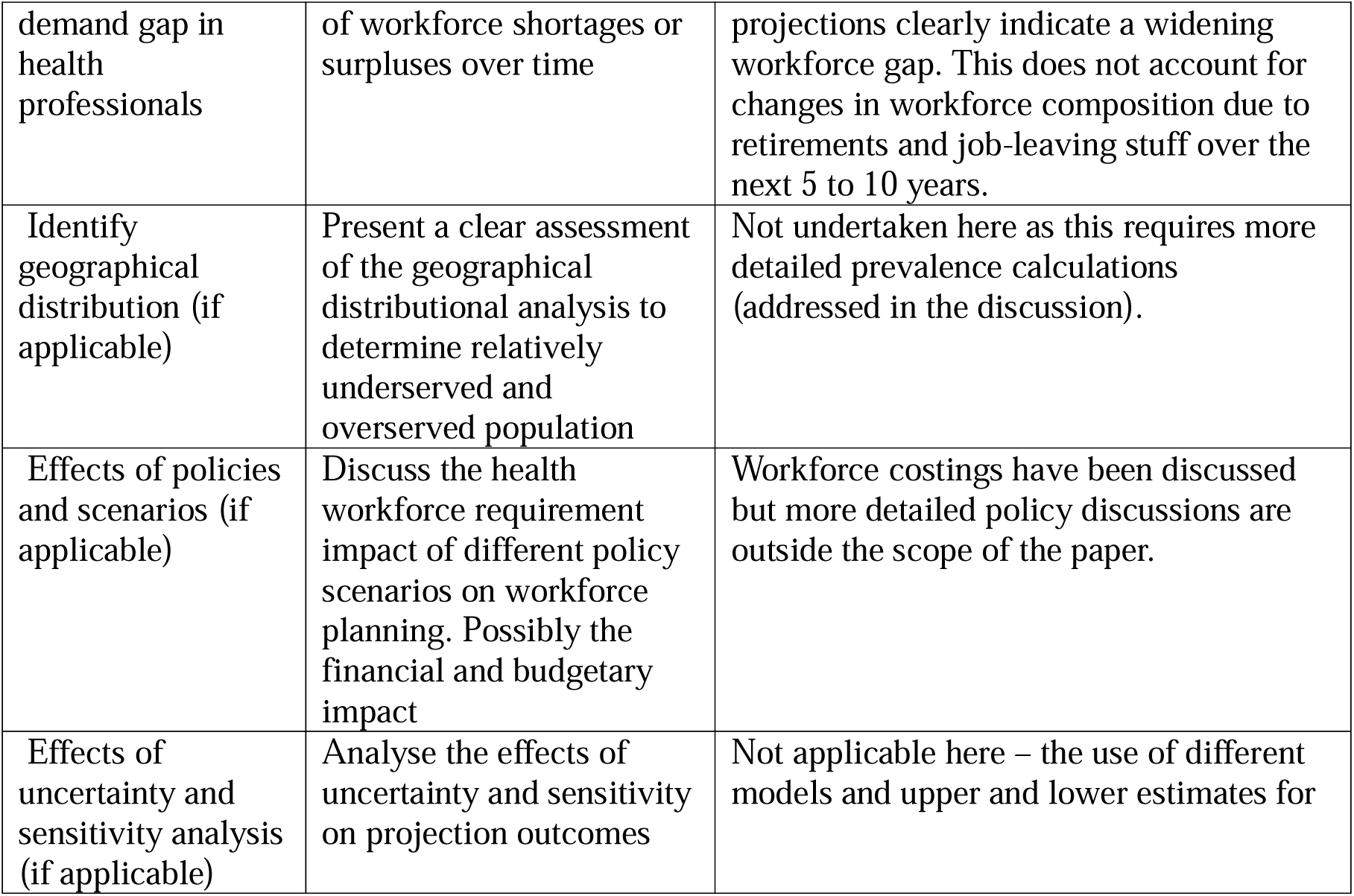

## Appendix 2 Topic guide for stakeholder groups

### Objectives

To understand the staffing needs for audiology services in the East of England based on views of Heads of Service using interviews, focus groups and surveys. The anticipated outcome of this research is the development of a data driven workforce planning tool for future NHS audiology staffing and training needs. In addition, the workforce planning tool and the underlying model can be used, with some adjustments, across all departments and specializations of the NHS in all four nations of the UK.

### Design

The focus groups will be comprised of Heads of Service who provide NHS audiology services for patients. A facilitator will explore the impact of experience as the service provider and challenges surrounding access and work in audiology. Broad themes

1. The impact of lived experience of hearing loss and service users of NHS audiology.
2. Lived experiences of service providers and staffing needs.
3. Suggestions for improving the experience of working in audiology or accessing services

| Main questions | Optional probe questions |
| --- | --- |
| What challenges have you encountered as a hearing care professional in providing timely services to your patients? | Where do you think, in your experiences, are most delays/barriers in service provision? |
| What do you consider a quality service to be? | What indicators of quality do you think would be useful? |
| Do you have any suggestions for the profession and professional bodies in improving the experience working as an audiology professional? |  |
| What changes do you anticipate in way audiology services will be provided in the next 10 years. |  |

## Appendix 3 BAA approved NHS East of England guidance for patient vs. non-patient-facing time according to band

Appendix 3 Title: BAA approved NHS East of England guidance for patient vs. non-patient-facing time according to band
| Staff band | Patient/non-Patient facing timings |
| --- | --- |
| Clinical AFC Band 3 | 90/10 |
| Clinical AFC Band 4 | 80/20 |
| Clinical AFC Band 5 | 80/20 |
| Clinical AFC Band 6 | 70/30 |
| Clinical AFC Band 7 | 65/35 |
| Clinical AFC Band 8a | 50/50 |
| Clinical AFC Band 8b | 50/50 |
| Clinical AFC Band 8c | 40/60 |
| Clinical AFC Band 8d | 30/70 |
| Clinical AFC Band 9 | 30/70 |
| Consultant Medical | 30/70 |
| Specialist Registrar | 30/70 |

## Appendix 4

Title: BAA approved NHS East of England guidance for role allocation of procedures for paediatric and adult audiology according to NHS Agenda For Change banding

| Paediatric Audiology Activities | Min band | Band 5 | Band 6 | Band 7 |
| --- | --- | --- | --- | --- |
| PTA 1 | 6 | 0% | 70% | 25% |
| VRA | 6 | 0% | 70% | 85% |
| Paediatric 1 person Fitting & Review | 6 | 0% | 0% | 70% |
| Paediatric 2 person Fitting & Review | 6 | 0% | 70% | 85% |
| Hearing aid repairs | 6 | 0% | 70% | 25% |
| Balance assessment | 8a | 0% | 0% | 0% |
| Vestibular rehabilitation (incl BPPV clinics) | 7 | 0% | 0% | 80% |
| Tinnitus appointments | 7 | 0% | 0% | 70% |
| On the day ENT hearing assessments | 5 | 33% | 47% | 20% |
| Electrophysiology | 7 | 0% | 70% | 30% |
| Wax removal | 7 | 0% | 60% | 40% |
| Auditory Processing Disorder (APD) | 7 | 0% | 0% | 70% |
| Bone Conduction Hearing Implant/MEI Assessments | 6 | 0% | 40% | 40% |
| Bone Conduction Hearing Implant/MEI fitting/activation/first year pathway | 6 | 0% | 40% | 40% |
| Bone Conduction Hearing Implant /MEI Device Upgrades or Reviews | 6 | 0% | 40% | 40% |

| Adult Audiology Activities | Min band | Band 2 | Band 3 | Band 4 | Band 5 | Band 6 | Band 7 |
| --- | --- | --- | --- | --- | --- | --- | --- |
| Hearing assessment (only) | 5 | 0% | 0% | 0% | 60% | 20% | 15% |
| Hearing assessment and fit | 5 | 0% | 0% | 0% | 60% | 20% | 15% |
| Hearing aid fitting (only) | 5 | 0% | 0% | 0% | 60% | 20% | 15% |
| Hearing aid review | 5 | 0% | 0% | 0% | 60% | 20% | 15% |
| Hearing aid repairs | 3 | 0% | 60% | 20% | 20% | 0% | 0% |
| Balance assessment | 7 | 0% | 0% | 0% | 0% | 0% | 80% |
| Vestibular rehabilitation (incl BPPV clinics) | 7 | 0% | 0% | 0% | 0% | 40% | 40% |
| Tinnitus appointments | 6 | 0% | 0% | 0% | 0% | 40% | 40% |
| On the day ENT hearing assessments | 5 | 0% | 0% | 0% | 60% | 20% | 15% |
| Electrophysiology | 7 | 0% | 0% | 0% | 0% | 0% | 80% |
| Wax removal | 4 | 0% | 0% | 0% | 0% | 40% | 40% |
| Auditory Processing Disorder (APD) | 7 | 0% | 0% | 0% | 0% | 0% | 80% |
| Bone Conduction Hearing Implant/MEI Assessments | 6 | 0% | 0% | 0% | 0% | 40% | 40% |
| Bone Conduction Hearing Implant/MEI fitting/activation/first year pathway | 6 | 0% | 0% | 0% | 0% | 40% | 40% |
| Bone Conduction Hearing Implant /MEI Device Upgrades or Reviews | 6 | 0% | 0% | 0% | 0% | 40% | 40% |

## Appendix 5 Reported Audiology Workforce by Band for England (2025)

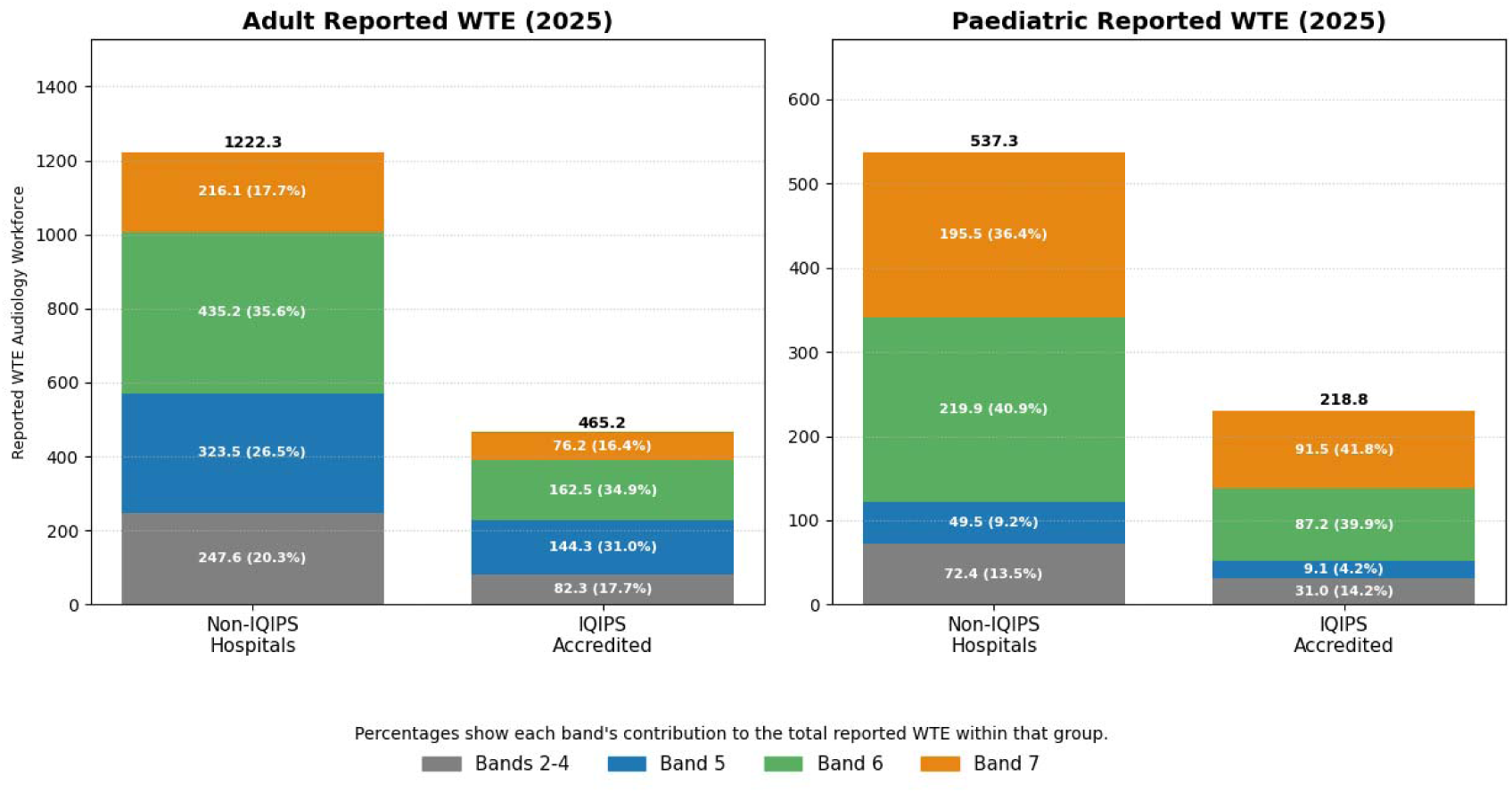

Caption: Current reported workforce is shown split by IQIPs accredited and non-accredited sites (colours show clinical banding -higher band numbers denote increasing in seniority and specialisation).

## Appendix 6 Adult WTE Workforce Estimations using Model 1.

| Band | 2025 | 2030 | % difference | 2035 | % difference |
| --- | --- | --- | --- | --- | --- |
| Bands 2-4 | 157.2<br>(158.5) | 162.7<br>(164.1) | 3.5%<br>(3.5%) | 167.8<br>(169.3) | 6.8%<br>(6.8%) |
| Band 5 | 303.0<br>(351.4) | 314.8<br>(365.1) | 3.9%<br>(3.9%) | 325.9<br>(378.1) | 7.6%<br>(7.6%) |
| Band 6 | 390.0<br>(458.6) | 405.2<br>(476.4) | 3.9%<br>(3.9%) | 419.6<br>(493.3) | 7.6%<br>(7.6%) |
| Band 7 | 196.9<br>(235.9) | 204.5<br>(245.1) | 3.9%<br>(3.9%) | 211.8<br>(253.8) | 7.6%<br>(7.6%) |
| Total | 1047.7<br>(1204.4) | 1087.2<br>(1250.6) | 3.77%<br>(4.41%) | 1125.2<br>(1294.4) | 7.40%<br>(7.47%) |

Caption: Projected workforce requirements for the data collection year (2025), 5-year (2030) and 10-year (2035) periods are shown alongside the percentage change from 2025 requirements for 2030 and 2035 across clinical banding (increasing in seniority and specialisation). Shown in () are the estimates when current waiting times are reduced to under 6 weeks (consistent with NHS recommendations on maximum waiting times for diagnostic services).

## Appendix 7 Paediatric Whole Time Equivalent Workforce Estimations (Model 1: Population Growth, Model 2: Steady State, Model 3: Mixed Model).

**Title: Paediatric Whole Time Equivalent Workforce Estimations (Model 1: Population Growth, Model 2: Steady State, Model 3: Mixed Model).**
| Band | 2025 | 2030<br>MODEL<br>1 | %<br>*difference* | 2030<br>MODEL<br>2 | %<br>*difference* | 2030<br>MODEL<br>3 | %<br>*difference* | 2035<br>MODEL<br>1 | %<br>*difference* | 2035<br>MODEL<br>2 | %<br>*difference* | 2035<br>MODEL<br>3 | %<br>*difference* |
| --- | --- | --- | --- | --- | --- | --- | --- | --- | --- | --- | --- | --- | --- |
| Bands<br>2-4 | 63.5<br>(63.5) | 64.0<br>(64.0) | 0.7%<br>(0.7%) | 64.0<br>(64.0) | 0.7%<br>(0.7%) | 64.0<br>(64.0) | 0.7%<br>(0.7%) | 64.5<br>(64.5) | 1.5%<br>(1.5%) | 64.5<br>(64.5) | 1.5%<br>(1.5%) | 64.5<br>(64.5) | 1.5%<br>(1.5%) |
| Band<br>5 | 18.7<br>(18.7) | 18.8<br>(18.8) | 0.7%<br>(0.7%) | 18.8<br>(18.8) | 0.7%<br>(0.7%) | 18.8<br>(18.8) | 0.7%<br>(0.7%) | 19.0<br>(19.0) | 1.5%<br>(1.5%) | 19.0<br>(19.0) | 1.5%<br>(1.5%) | 19.0<br>(19.0) | 1.5%<br>(1.5%) |
| Band<br>6 | 255.5<br>(424.3) | 246.0<br>(408.4) | -3.7%<br>(-3.7%) | 344.3<br>(571.8) | 34.8%<br>(34.8%) | 329.2<br>(546.6) | 28.8%<br>(28.8%) | 239.5<br>(387.0) | -6.3%<br>(-8.8%) | 464.4<br>(771.0) | 81.7%<br>(81.7%) | 428.9<br>(712.2) | 67.9%<br>(67.9%) |
| Band<br>7 | 288.6<br>(466.3) | 277.8<br>(448.9) | -3.7%<br>(-3.7%) | 388.9<br>(628.4) | 34.8%<br>(34.8%) | 371.8<br>(600.7) | 28.8%<br>(28.8%) | 270.5<br>(437.1) | -6.3%<br>(-6.3%) | 524.5<br>(847.4) | 81.7%<br>(81.7%) | 484.5<br>(782.7) | 67.9%<br>(67.9%) |
| Total | 626.3<br>(972.8) | 606.6<br>(940.1) | -3.15%<br>(-3.36%) | 816<br>(1283) | 30.29%<br>(31.89%) | 783.8<br>(1230.1) | 25.15%<br>(26.45%) | 593.5<br>(907.6) | -5.24%<br>(-6.70%) | 1072.4<br>(1701.9) | 71.23%<br>(74.95%) | 996.9<br>(1578.4) | 59.17%<br>(62.25%) |

Caption: Projected workforce requirements for the data collection year (2025), 5-year (2030) and 10-year (2035) periods are shown alongside the percentage change from 2025 requirements for 2030 and 2035 across clinical banding (increasing in seniority and specialisation). Shown in () are the estimates when current waiting times are reduced to under 6 weeks (consistent with NHS recommendations on maximum waiting times for diagnostic services).

